# Cross-cohort validation of EEG biomarkers using a pre-registered three-criterion framework: a demonstration in amyotrophic lateral sclerosis

**DOI:** 10.64898/2026.11.10.26367741

**Authors:** Belay Tadesse Gebregergis, Haben Girmay Yhdego, Tewolde Teklu

## Abstract

**Background:** Despite decades of EEG biomarker development for amyotrophic lateral sclerosis (ALS), no candidate has progressed to clinical deployment, partly because cross-cohort validation is rare. We aimed to develop and demonstrate a pre-registered three-criterion screening framework to identify which candidate EEG features show the combined properties required for biomarker translation: cross-sectional discrimination, within-subject reliability, and cross-cohort effect consistency.

**Methods:** We pre-registered a validation framework adapted from FDA-NIH BEST principles and applied it to 18 EEG features across three independent cohorts: EEGET-ALS (six ALS patients, 170 healthy controls; home recordings over 17 days), UCL/Penn State (eight ALS patients; clinic recordings over 1-2 months), and a healthy adult test-retest reference (60 participants, three sessions over one month). Features were required to meet three pre-registered criteria: (1) cross-sectional discrimination (Cohen’s d > 0.5), (2) within-subject reliability (ICC(3,1) > 0.6 in both ALS cohorts independently), and (3) cross-cohort effect direction consistency. Cross-cohort permutation testing (1000 iterations) and subject-level bootstrap resampling (500 iterations) were applied to assess robustness.

**Results:** Three features met all three pre-registered criteria: gamma power, theta power, and spectral entropy. Cross-cohort permutation testing showed that no permutation, out of 1000, produced any feature in the three-cohort reliability intersection (Monte Carlo p = 0.001). Bootstrap resampling indicated that the composition of the selected set was sensitive to sample composition (median count 1, 95% CI [0, 4]), with gamma power (67% inclusion), spectral entropy (43%), and theta power (32%) the most stable features at a relaxed reliability threshold (ICC > 0.5); at the primary pre-registered threshold (ICC > 0.6), the corresponding inclusion probabilities are lower (57%, 27%, and 19%, respectively). A principal methodological finding was that cross-cohort differences in feature reliability were large relative to the apparent disease-associated signal, most clearly demonstrated by relative beta power (ICC = 0.09 in a home-recorded cohort versus 0.93 in a clinic-recorded cohort); because recording protocol, hardware, setting, population, and preprocessing all differ jointly between cohorts in this dataset, this is evidence of cohort-associated variability that plausibly includes a protocol contribution, not a separately identified, isolated protocol effect.

**Conclusions:** This study provides a pre-registered, multi-cohort EEG biomarker screening framework for ALS with explicit bootstrap-quantified uncertainty. Gamma power, spectral entropy, and theta power are identified as priority candidates for prospective validation. A recurring methodological observation is that protocol-related cohort differences appear to contribute more to cross-cohort variability than disease-related effects within this dataset; because protocol is confounded with cohort, equipment, and preprocessing, this pattern is descriptive rather than causal. The observation nonetheless has direct implications for the design of multi-site EEG biomarker studies in neural engineering.

## 1. Introduction

Amyotrophic lateral sclerosis (ALS) is a progressive motor neuron disease with a median survival of approximately 3-5 years after symptom onset [1, 2]. Clinical trials in ALS remain limited by the absence of validated biomarkers that can sensitively track disease progression and detect treatment effects with feasible sample sizes [3].

Electroencephalography (EEG) is a promising candidate biomarker modality for ALS because it is non-invasive, relatively inexpensive, and compatible with repeated or decentralised acquisition. Resting-state EEG abnormalities in ALS have been reported across multiple feature domains, including alterations in spectral power, oscillatory organisation, and functional connectivity linked to motor and cognitive phenotypes [4–9]. However, most ALS-EEG studies remain cross-sectional and single-cohort, with limited evaluation of whether candidate features generalise across acquisition settings or show stable within-subject measurement properties [3, 4].

This gap presents a major challenge for biomarker translation. The FDA-NIH BEST framework emphasises both clinical validity (discrimination of relevant clinical states) and analytical validity (reliable and reproducible measurement) [10], while EEG-methodology studies consistently show that many electrophysiological features are sensitive to acquisition protocols, recording contexts, and sample characteristics [11]. Current ALS-EEG studies typically rely on single cohorts, cross-sectional designs, and post-hoc analytic choices, none of which establishes that a candidate biomarker is reliable across recording contexts or robust to analytic flexibility. For neural engineering applications, EEG biomarkers intended for clinical or decentralised deployment must show not only cross-sectional discrimination but also reproducibility across sessions, cohorts, and recording environments.

To address this need, we present a pre-registered multi-cohort EEG biomarker framework in ALS that jointly evaluates candidate features along three complementary dimensions: (1) discrimination between ALS and healthy controls, (2) within-subject reliability within independent ALS cohorts, and (3) consistency of disease-related effect direction across cohorts [6, 11, 12]. Rather than claiming biomarker validation, our objective was to identify EEG features that show the strongest combined evidence of discriminative utility, measurement stability, and cross-cohort reproducibility, while highlighting feature classes disproportionately influenced by protocol-dependent variability. Such evaluation is essential for the development of scalable EEG biomarkers suitable for longitudinal and translational neural engineering applications in ALS.

## 2. Methods

### 2.1 Pre-registration and design

The validation framework (Fig. 1) was pre-registered on the Open Science Framework (OSF, doi:10.17605/OSF.IO/4K32S) before the final analyses. All thresholds, the 18-feature panel, three-cohort design, MAD-normalised preprocessing, and bootstrap/permutation defence procedures were specified before computing the results. Two pre-specified clarifications were added during data inspection but before final results: (1) restriction of UCL trials to the first 160 per subject to enforce the dataset’s stated 4-session x 40-trial design, and (2) exclusion of two delta-band features after cross-cohort spectral inspection revealed an order-of-magnitude reduction in UCL delta-band power consistent with undocumented preprocessing in that band.

**Figure 1.**
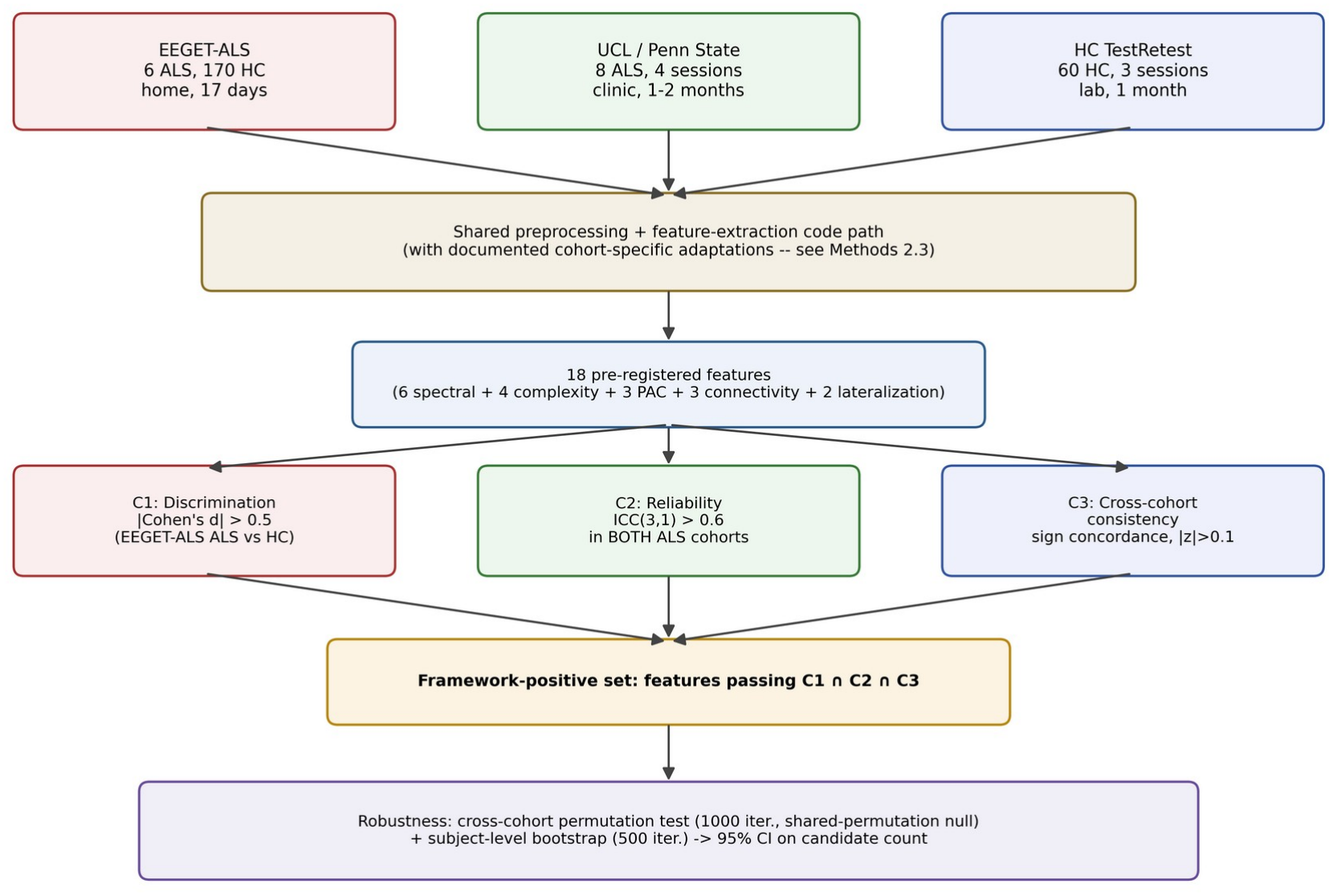
Schematic of the pre-registered three-cohort EEG biomarker validation framework. Three independent cohorts (EEGET-ALS, UCL/Penn State, HC TestRetest) are processed through a shared preprocessing and feature-extraction code path, with the documented cohort-specific adaptations described in Methods 2.3. The 18 pre-registered features are evaluated against three primary criteria (C1 discrimination, C2 reliability in both ALS cohorts, C3 cross-cohort consistency). Features passing all three constitute the framework-positive set. Cross-cohort permutation testing and subject-level bootstrap assess framework robustness and provide a confidence interval on the priority candidate count. For narrative simplicity, the three features that pass the full three-criterion framework are highlighted as selected candidates for prospective validation; the full ranked bootstrap-inclusion distribution is shown in figure 3.

### 2.2 Cohorts

Cohort characteristics are summarised in (Table 1).

**Table 1.**
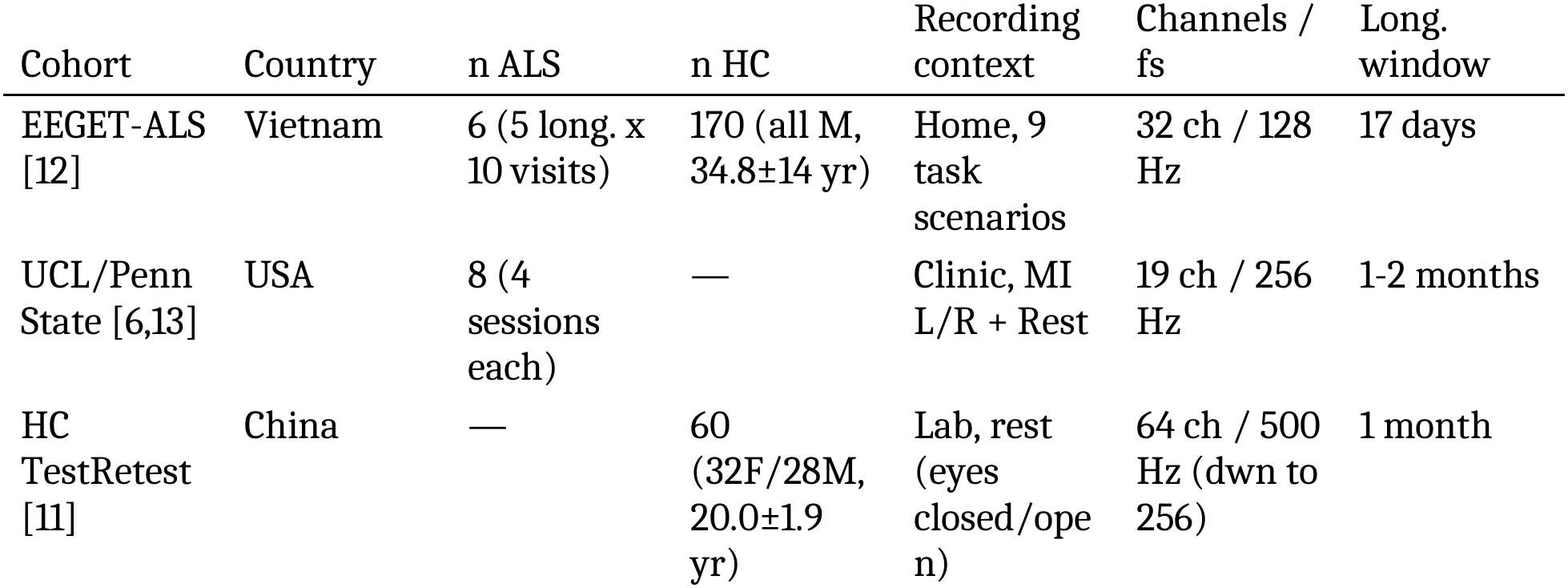
Cohort characteristics.

#### EEGET-ALS [12]

The Vietnamese cohort contained six ALS patients (three males, three females; mean age 56 years, range 37-77) and 170 healthy controls (all males; mean age 34.8 years, range 18-82). Recordings were home-based across nine task scenarios using a 32-channel EEG at 128 Hz. Five ALS patients contributed longitudinal data across 10 visits spanning 17 days, yielding 50 visit-level recordings for within-subject reliability analysis. Total recordings included: 1988. Per-task discrimination and reliability values are shown in (Fig. S1).

#### UCL/Penn State [6, 13]

Eight patients with ALS underwent four sessions each across 1-2 months. Each session contained motor imagery (left and right) and rest trials. Recordings were trial-based and clinic-collected at 256 Hz with 19 channels (three EOG channels excluded). The first 160 trials per subject were used (four sessions x 40 trials).

#### HC Test Retest [11]

Sixty healthy adult participants (32 female, 28 males; mean age 20.0 years, range 18-28) from Southwest University, Chongqing, China. Three sessions per participant: two within-day and one one-month follow-up. Eyes-closed and eyes-open resting states (5 minutes each). 64-channel recording at 500 Hz; 19 channels matching the UCL set were selected, and data were re-referenced to the common average.

### 2.3 Preprocessing

A common preprocessing pipeline (the same functions and parameter choices) was applied across all three cohorts, adapted for each cohort’s native recording structure; the two most consequential adaptations – epoching strategy and notch frequency – are noted below, and are not minor implementation details: UCL’s trial-segmented recordings are analysed per-trial with no further epoching, while EEGET-ALS and HC Test-Retest, which are continuous recordings, are cut into fixed-length epochs, so “common pipeline” should be read as shared code and parameters rather than literally identical processing of the raw signal. Each recording was MAD-normalised per channel (subtracting the median and dividing by the median absolute deviation) to neutralize cross-cohort differences in nominal recording units. Recordings were bandpass-filtered (1-45 Hz, zero-phase Butterworth), notch-filtered at 50 Hz (Vietnam and China) or 60 Hz (UCL/US), and re-referenced to the common average. HC Test-Retest data were down sampled from 500 to 256 Hz. Channels exceeding ±30 MAD-normalised units or with point-to-point gradients exceeding ±5 were flagged and replaced with the sample mean of the remaining channels; we did not use spherical-spline interpolation, since accurate sensor coordinates are not reliably available from the low-density Emotiv EPOC+ exports used by EEGET-ALS. EEGET-ALS and HC Test-Retest recordings are continuous and were cut into 2-second epochs with 50% overlap, with bad epochs excluded; UCL recordings are natively trial-segmented (discrete motor-imagery/rest trials), so each trial was used directly as the analysis epoch without further re-epoching. Features were computed per epoch/trial and aggregated to the recording level using the mean. Median aggregation was the pre-registered intent; the results reported here use mean aggregation instead. This is a deviation from the pre-registration, disclosed here rather than silently substituted: mean aggregation is the aggregator that was actually run across all three cohorts. Re-running the full extraction pipeline with median aggregation – three independent extraction pipelines against raw EEG – was not performed before reporting these exploratory screening results. Whether the framework-positive set is sensitive to this choice has not been tested and is noted as an open limitation (see Limitations), not one this manuscript claims to have reason to discount.

### 2.4 Feature panel

Twenty candidate features compiled from the ALS EEG biomarker literature were pre-registered, organised into five families; per-feature literature provenance is documented in (Table S1). After pilot inspection revealed an order-of-magnitude reduction in UCL relative delta-band power consistent with undocumented preprocessing (Fig. S2), spec_delta and mvl_delta_gamma were excluded from the primary panel, reducing it to 18 features. Both excluded features are reported in (Table S2).

The 18 primary features were:

Spectral (6): relative power in theta (4-8 Hz) [4, 9, 7, 14, 15], alpha (8-13 Hz) [16, 14, 17], beta (13-30 Hz) [18, 5, 19, 20], and gamma (30-45 Hz) [18, 7, 21, 19, 9]; peak alpha frequency [24, 25]; spectral entropy [26].

Complexity (4): Hjorth mobility [27], Hjorth complexity [27], sample entropy [26], and permutation entropy [26].

Phase-amplitude coupling (3): mean vector length for alpha-gamma [23] and theta-gamma [22]; modulation index for alpha-gamma [23].

Connectivity (3): C3-C4 alpha coherence [18, 7, 9], C3-C4 beta coherence [18, 5, 7], mean alpha coherence [18, 7].

Lateralisation (2): C3-C4 alpha [6, 17, 28] and C3-C4 beta lateralisation [6].

(6 + 4 + 3 + 3 + 2 = 18. The excluded-from-primary delta-band features, `spec_delta` and `mvl_delta_gamma`, belonged to the Spectral and PAC families respectively, which is why the original twenty-feature pre-registered panel had one more member in each of those two families.)

### 2.5 Three primary criteria

#### Criterion 1 (Discrimination)

Cohen’s d between ALS (n = 6) and HC (n = 170) at the EEGET-ALS subject level. Threshold: |d| > 0.5 [29]. Bootstrap 95% CI on d via subject-level resampling.

#### Criterion 2 (Within-subject reliability)

ICC(3,1) computed independently for each ALS cohort: in EEGET-ALS across 10 visits of the five longitudinal ALS subjects; in UCL across four sessions of all eight subjects. Threshold: ICC > 0.6 in both cohorts independently [30]. This threshold corresponds to the lower bound of the moderate reliability range and was selected as appropriate for exploratory screening rather than for clinical deployment (for which ICC > 0.75 or 0.9 would be more conventional). ICC(3,1) (two-way mixed-effects, single-rater, absolute agreement, Case 3A of [33], building on the two-way random/mixed framework of [31]) was chosen because (a) visits are not exchangeable random selections from a broader population, ruling out ICC(2,1), and (b) features that systematically drift across visits should not be counted as reliable, ruling out consistency-only formulations. This choice is conservative. Concretely, for each feature and cohort the subject x visit(or session) value matrix was decomposed by two-way ANOVA into subject, occasion, and residual mean squares (MSR, MSC, MSE); ICC(3,1) absolute agreement was computed as (MSR - MSE) / (MSR + (k-1)*MSE + (k/n)*(MSC - MSE)), where n is the number of subjects and k the number of occasions. This requires a complete (no missing visits) subject x occasion design, which held for all three cohorts as extracted.

#### Criterion 3 (Cross-cohort effect consistency)

For each feature, the mean ALS score in each cohort was z-scored against the EEGET-ALS HC distribution. A feature passed criterion 3 if both z-scores had the same sign and |z| > 0.1. The |z| > 0.1 floor excluded features whose deviation from healthy controls was so small that sign concordance was statistically uninformative. As a sensitivity check, removing this floor changed the number of features passing criterion 3 by at most ±1 and left the three-criterion intersection unchanged (Table S3). Because both ALS cohorts are standardised against the same EEGET-ALS healthy-control reference rather than each having its own independent control group, Criterion 3 measures directional concordance relative to a shared reference distribution, not a fully independent replication of the ALS-versus-control effect in each cohort.

### 2.6 Bootstrap and permutation defence

#### Cross-cohort permutation test (1000 iterations)

For each cohort, the observed subject x occasion value matrix (n subjects x k visits/sessions) was held fixed in shape; within each occasion (column), a single random subject relabeling was drawn per cohort per iteration and applied identically to every one of the 18 primary features (all of which share an identical, complete subject x occasion pivot within a given cohort). Applying one shared relabeling across features, rather than an independent one per feature, preserves cross-feature covariance in the resulting null distribution – necessary because the test statistic (the count of features simultaneously in the three-cohort reliability intersection) is a joint, not per-feature, quantity. This destroys within-subject (row) coherence – the signal the null hypothesis says should not exist – while preserving each occasion’s marginal distribution and the cohort’s exact subject/occasion balance. ICC(3,1) was recomputed on the permuted matrix in each cohort and the number of features in the three-cohort reliability intersection (ICC > 0.6 in all three cohorts simultaneously) was counted. The test asks: if within-subject values carried no subject-specific signal, how often would any feature still pass the three-cohort intersection by chance? The p-value was computed with the standard Monte Carlo estimator, p = (1 + #{null count >= observed count}) / (1 + 1000), which is bounded away from exactly zero by construction. The number of iterations (1000) gives a minimum resolvable p-value of 0.001; increasing the number of iterations to 5000 or 10000 would refine the tail estimate but not change the qualitative conclusion.

#### Subject-level bootstrap (500 iterations)

Within each cohort, subjects were resampled with replacement at the original cohort size. All three criteria were re-evaluated for each resample. The proportion of bootstrap iterations in which each feature appeared in the priority candidate set is its bootstrap inclusion probability.

### 2.7 Variance decomposition

For each feature in each cohort, total visit-level variance was decomposed into between-subject and within-subject components using a one-way ANOVA. The between-subject proportion (signal) was bounded between zero and one. This decomposition complemented ICC by exposing features that fail to be reliable due to small between-subject differences versus large within-subject noise.

## 3. Results

### 3.1 Cohort characteristics and feature extraction

Of 1989 EEGET-ALS recordings, 1988 (99.95%) passed quality control. All 96 UCL session-level recordings (eight subjects x four sessions x three trial classes) were retained. All 360 HC TestRetest recordings (60 subjects x three sessions x two conditions) were retained. The final feature matrix consisted of 1988 EEGET-ALS, 96 UCL, and 360 HC rows, each containing an 18-feature primary panel. Full cohort demographics and recording parameters are summarised in (Table 1).

### 3.2 Three features emerge as framework-positive candidates

By applying the full pre-registered three-criterion framework at the moderate threshold (ICC > 0.6), three features emerged as framework-positive: gamma power, theta power, and spectral entropy (Table 2). These features cleared cross-sectional discrimination (|d| > 0.5), within-subject reliability (ICC > 0.6 in both ALS cohorts independently), and cross-cohort effect-direction consistency. Of the 18 primary features, 9 passed criterion 1, six passed criterion 2, and seven passed criterion 3 individually; the three-feature intersection reflected substantial winnowing by the joint requirements. The full ranked scorecard with confidence intervals is provided in (Table S4).

**Table 2.** Per-feature scorecard for 18 primary features (point estimates; full 95% confidence intervals for Cohen’s d and each ICC are in Table S4). Cohen’s d (sample-size-weighted pooled SD) computed on subject-averaged EEGET-ALS data (ALS vs HC). ICC EG = two-way mixed-effects ICC(3,1), absolute agreement, in EEGET-ALS; ICC UCL = same in UCL; ICC HC = same in HC TestRetest. C1 = passes discrimination (|d| > 0.5). C2 = passes reliability in both ALS cohorts (ICC > 0.6). C3 = passes cross-cohort sign concordance with |z| > 0.1 floor. Pass = criteria passed out of 3. Bootstrap % shown at ICC > 0.5 threshold. Feature names are given in prose form here for table legibility; the corresponding code/data-column identifiers (e.g. “Gamma power” = `spec_gamma`) are listed in Section 2.4 and in `results/c1_discrimination.csv` / `results/c2_reliability.csv`. All pass/fail decisions (C1-C3, Pass) use unrounded values; two-decimal display can make a borderline case look ambiguous – e.g. theta power’s ICC EG rounds to 0.60 but the unrounded value (0.6049) clears the ICC > 0.6 threshold.

| Feature | Family | d | ICC EG | ICC UCL | ICC HC | C1 | C2 | C3 | Pass | Boot % |
|---|---|---|---|---|---|---|---|---|---|---|
| Gamma power | Spectral | +0.95 | 0.67 | 0.84 | 0.60 | Y | Y | Y | 3/3 | 67% |
| Theta power | Spectral | -0.87 | 0.60 | 0.82 | 0.74 | Y | Y | Y | 3/3 | 32% |
| Spectral entropy | Spectral | +0.75 | 0.64 | 0.71 | 0.68 | Y | Y | Y | 3/3 | 43% |
| Alpha peak freq. | Spectral | +0.13 | 0.71 | 0.84 | 0.85 | - | Y | Y | 2/3 | 14% |
| Alpha power | Spectral | -0.29 | 0.66 | 0.88 | 0.79 | - | Y | - | 1/3 | 1% |
| Beta power | Spectral | +0.79 | 0.09 | 0.93 | 0.76 | Y | - | Y | 2/3 | 1% |
| Permutation entropy | Complexity | +0.41 | 0.65 | 0.55 | 0.77 | - | - | - | 0/3 | 0% |
| Hjorth mobility | Complexity | +0.80 | 0.42 | 0.82 | 0.65 | Y | - | - | 1/3 | 0% |
| Hjorth complexity | Complexity | -0.67 | 0.49 | 0.87 | 0.59 | Y | - | Y | 2/3 | 30% |
| Sample entropy | Complexity | +0.11 | 0.51 | 0.60 | 0.77 | - | - | - | 0/3 | 3% |
| PAC a-g (MVL) | PAC | -0.89 | 0.31 | 0.53 | 0.42 | Y | - | - | 1/3 | 0% |
| PAC a-g (MI)* | PAC | +0.13 | -0.03 | -0.01 | 0.70 | - | - | Y | 1/3 | 0% |
| PAC t-g (MVL) | PAC | -0.88 | 0.26 | 0.69 | 0.44 | Y | - | - | 1/3 | 0% |
| C3-C4 a coherence | Connect. | -0.07 | 0.27 | 0.73 | 0.44 | - | - | - | 0/3 | 0% |
| C3-C4 b coherence | Connect. | +0.72 | 0.03 | 0.83 | 0.42 | Y | - | - | 1/3 | 0% |
| Mean a coherence | Connect. | +0.19 | 0.72 | 0.73 | 0.44 | - | Y | - | 1/3 | 0% |
| C3-C4 a lateraliz. | Lateral. | -0.31 | 0.17 | 0.33 | 0.28 | - | - | - | 0/3 | 0% |
| C3-C4 b lateraliz. | Lateral. | -0.42 | 0.21 | 0.04 | 0.33 | - | - | - | 0/3 | 0% |

\* PAC alpha-gamma (MI)’s effect size is near zero and its bootstrap CI upper bound is inflated by a small number of resamples with a near-zero pooled SD denominator; it does not affect any criterion-passing conclusion, since this feature fails C1 regardless.

A related four-feature reliability intersection (ICC > 0.6 in all three cohorts, ignoring discrimination and concordance criteria) contained alpha peak frequency, alpha power, theta power, and spectral entropy. Of these, two (theta power and spectral entropy) also passed the full three-criterion framework; the other two (alpha peak frequency and alpha power) failed criterion 1 (|d| = 0.13 and 0.29, respectively). This intersection was the unit of analysis for cross-cohort permutation testing.

### 3.3 Cross-cohort permutation testing supports framework validity

The cross-cohort permutation test (Fig. 2) rejected the null hypothesis. Across 1000 permutations, each of which independently shuffled feature values across subjects within each occasion of each cohort (preserving the subject/occasion balance while destroying within-subject coherence), zero permutations produced any feature in the three-cohort reliability intersection (null mean = 0.000, null max = 0; Monte Carlo p = 0.001, using the (b+1)/(B+1) estimator). This indicates that the observed four-feature reliability intersection is unlikely to have arisen under a null model with no genuine within-subject signal. Because the permutation preserves each cohort’s exact subject/occasion balance and each occasion’s marginal distribution but eliminates subject-specific coherence, this result shows that the intersection depends on coherent subject-level signals within each cohort; it does not by itself rule out cohort-level confounds.

**Figure 2.**
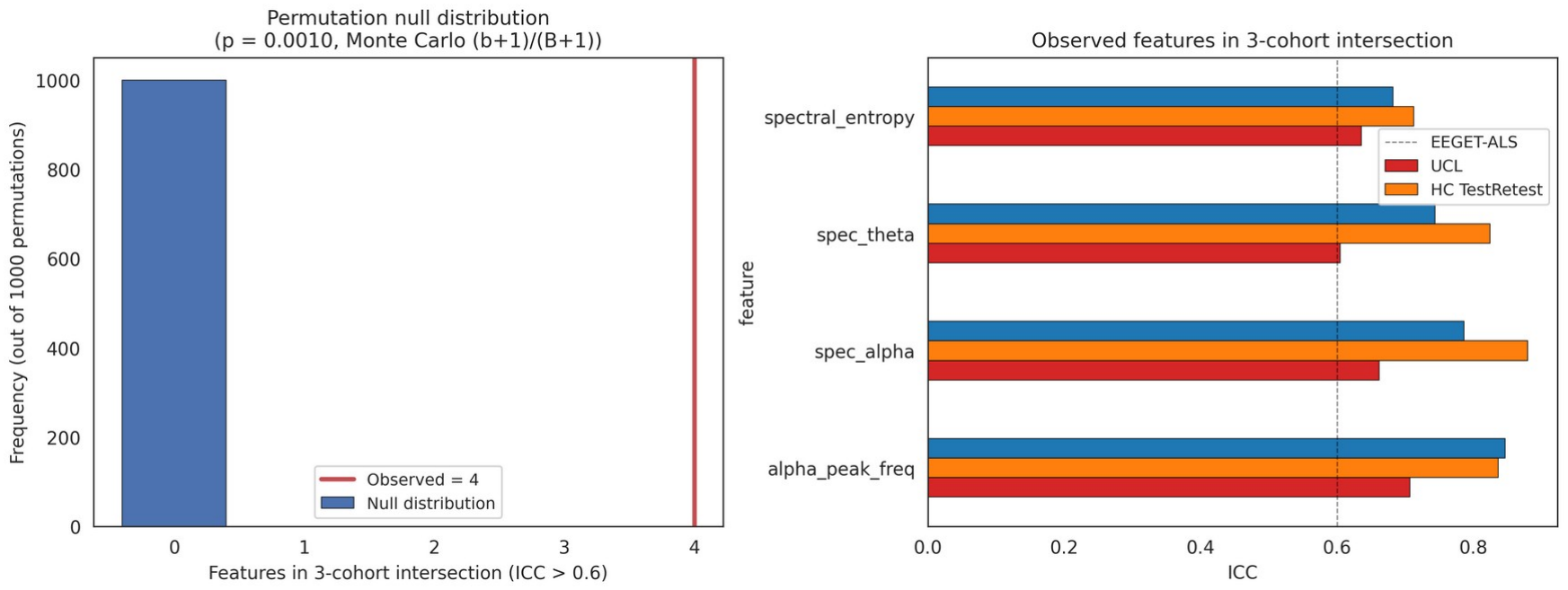
Cross-cohort permutation test results. (A) Null distribution of the number of features appearing in the three-cohort reliability intersection under within-occasion value permutation in each cohort (1000 iterations). All 1000 permutations produced zero features in the intersection. The observed result (4 features) is therefore beyond every value in the null distribution (Monte Carlo p = 0.001). (B) The four observed features in the three-cohort intersection at ICC > 0.6: alpha_peak_freq, spec_alpha, spec_theta, spectral_entropy.

### 3.4 Bootstrap stability reveals sample-size-driven uncertainty

Despite the significance of the permutation test, the precise composition of the selected set was sensitive to the subjects included (Fig. 3). Across 500 subject-level bootstrap resamples (drawn coherently: within a given resample, the same resampled EEGET-ALS subjects are used for both the discrimination and reliability criteria, rather than resampling each criterion’s subject set independently), the median count of selected features was 1 (95% CI [0, 4]) at the moderate threshold (ICC > 0.6) and 2 (95% CI [0, 4]) at the relaxed threshold (ICC > 0.5). At ICC > 0.5, gamma power appeared in the selected set in 67% of bootstrap resamples, spectral entropy in 43%, theta power in 32%, Hjorth complexity in 30.0%, and alpha peak frequency in 14%. All remaining features appeared in fewer than 5% of resamples. For the purpose of narrative simplicity we highlight the three features that also pass the full three-criterion framework at the primary ICC > 0.6 threshold (gamma power, spectral entropy, theta power) as the selected candidates for prospective validation. Hjorth complexity’s bootstrap inclusion probability lands exactly at the 30% pragmatic cutoff used in Fig. 3, but it does not pass Criterion 2 at the primary threshold (Table 2) and is therefore not included among the selected candidates; it is flagged here rather than silently omitted, since its inclusion probability is a boundary case rather than a clearly negative one.

**Figure 3.**
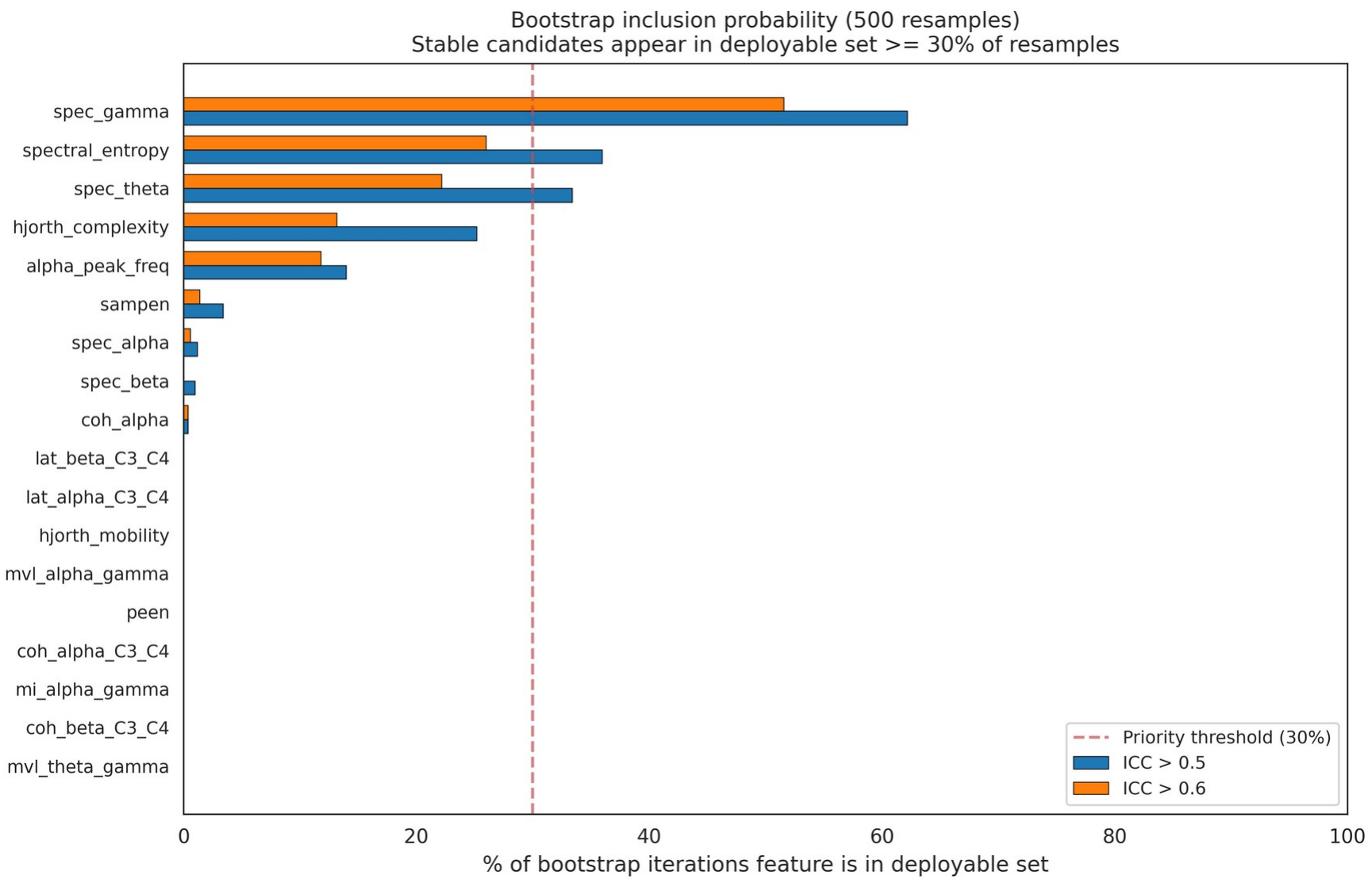
Per-feature bootstrap inclusion probability across 500 subject-level resamples. Each bar shows the proportion of bootstrap resamples in which a given feature appeared in the selected set, computed at both the moderate (ICC > 0.6, orange) and relaxed (ICC > 0.5, blue) reliability thresholds. Features are sorted by inclusion probability at ICC > 0.5. The dashed vertical line marks the 30% pragmatic reporting cutoff. Three features (gamma power, spectral entropy, theta power) clearly exceed this cutoff and are the most consistently recovered across resamples; a fourth (Hjorth complexity) lands exactly at the cutoff (30.0%) but does not pass Criterion 2 at the primary threshold and is not counted among the selected candidates (see Results 3.2).

### 3.5 Protocol-related differences dominate cross-cohort reliability variability

A three-cohort reliability comparison (Fig. 4) revealed an unexpected dissociation. The clearest case was relative beta power: ICC = 0.09 in EEGET-ALS (home recordings, 17-day window) but ICC = 0.93 in UCL (clinic recordings, 1-2-month window) and ICC = 0.76 in HC TestRetest (clinic recordings, 1-month window). This dissociation is unlikely to be fully explained by disease severity (UCL patients were on average more functional than EEGET-ALS patients) or to the longitudinal window (UCL window was longer, which should have increased rather than decreased within-subject variance). Variance decomposition (Fig. 5) confirmed that in EEGET-ALS only 15.6% of relative beta power’s total variance was between-subject, compared to 93.7% in UCL and 83.7% in HC; a 78-percentage-point gap between EEGET-ALS and UCL for the same feature computed by the same algorithm on the same channel set. Across the full 18-feature panel, 16 of 18 features showed a between-subject proportion gap of more than 20 percentage points between EEGET-ALS and either UCL or HC (11 features for the EEGET-ALS/UCL comparison alone, 15 for EEGET-ALS/HC); only coh_alpha, spectral_entropy, and peen stayed within 20 percentage points of EEGET-ALS in both other cohorts.

**Figure 4.**
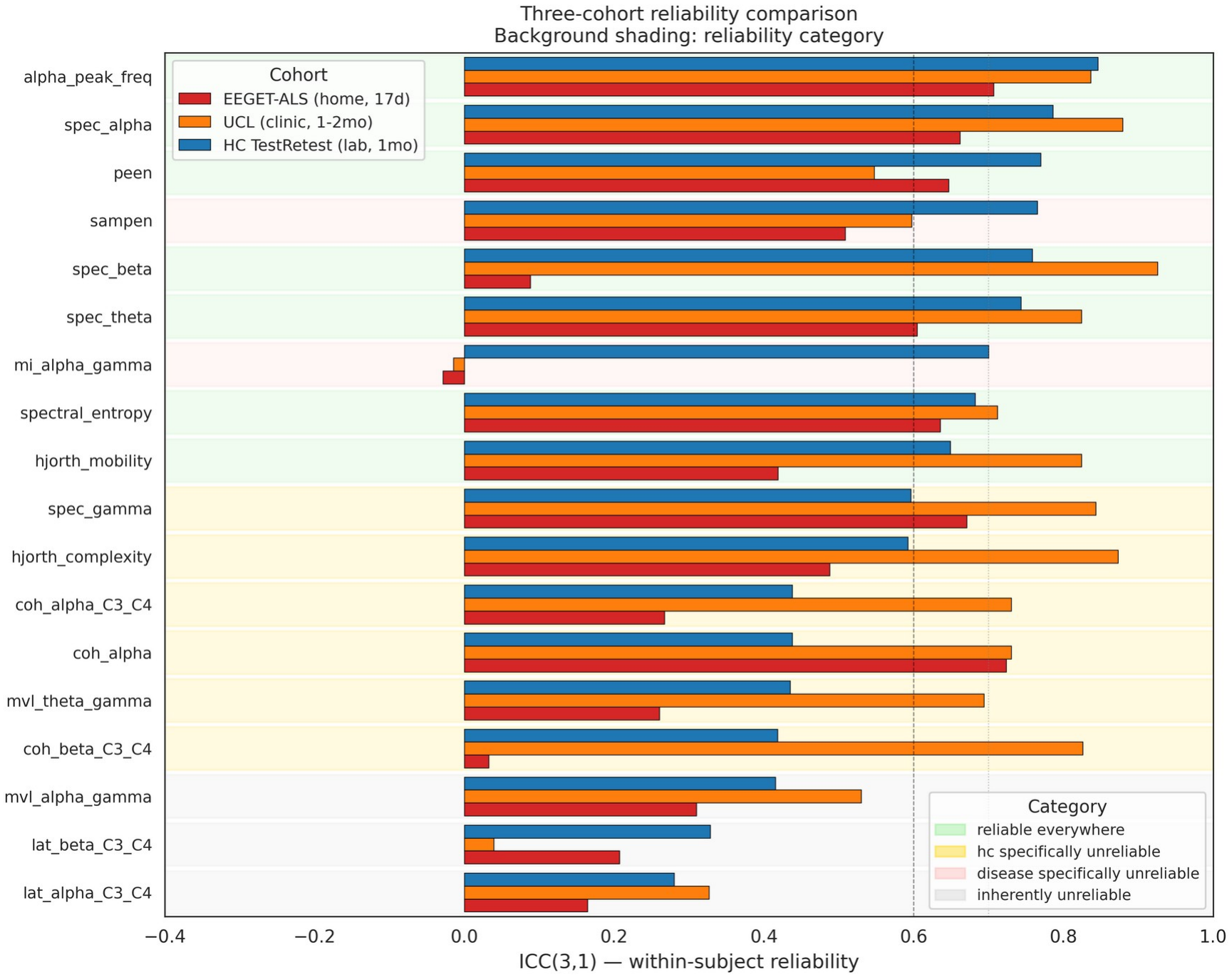
Three-cohort reliability comparison. ICC(3,1) for each feature in EEGET-ALS, UCL/Penn State, and HC TestRetest. Features sorted by HC ICC. Background shading indicates reliability category: green (reliable in all 3 cohorts), yellow (HC-specifically unreliable, i.e., reliable in ALS cohorts but not HC), pink (disease-specifically unreliable), gray (inherently unreliable). Note that spec_beta shows ICC = 0.09 in EEGET-ALS versus ICC = 0.93 in UCL and 0.76 in HC – a cohort-associated difference potentially arising from protocol, hardware, setting, population, or preprocessing differences between cohorts, not a separately identified “protocol effect” (see Discussion).

**Figure 5.**
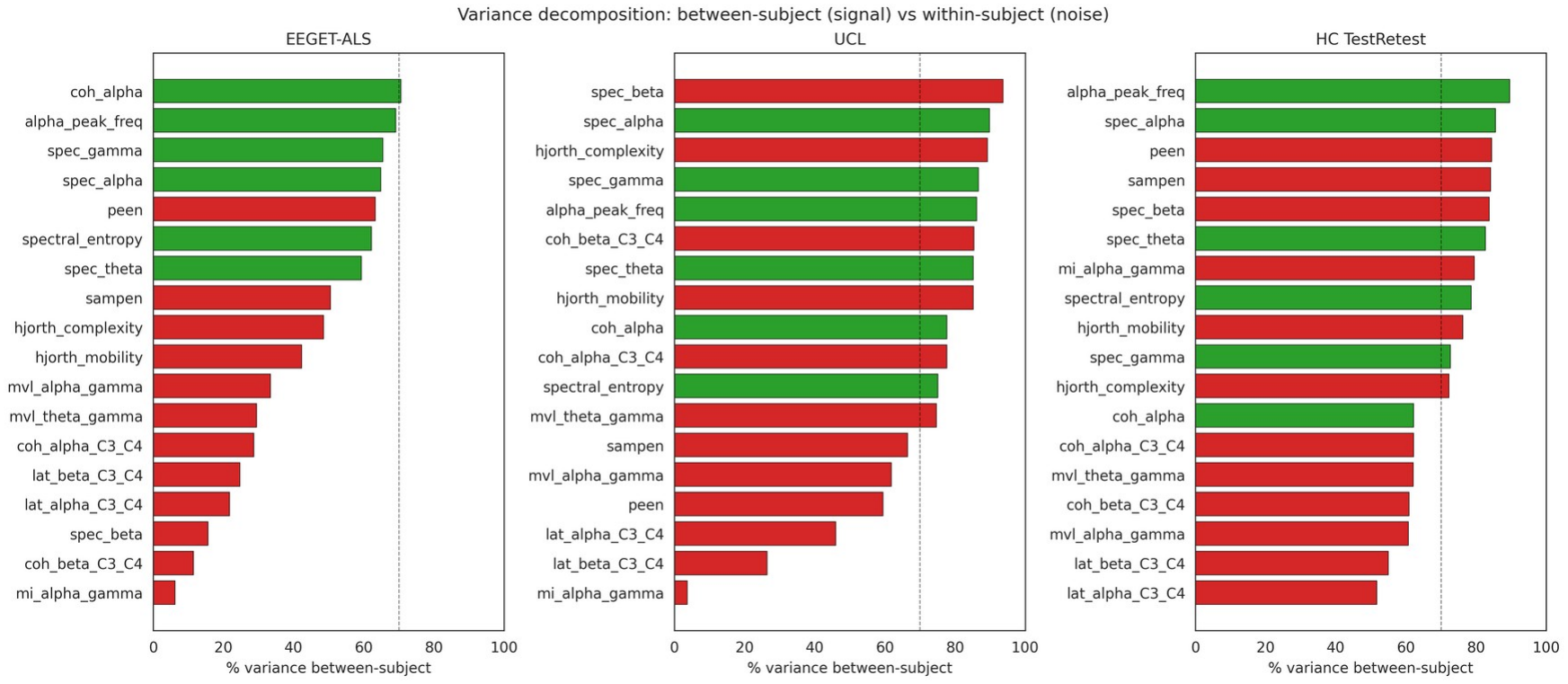
Variance decomposition: cross-cohort differences in between-subject variance structure. Fraction of total variance attributable to between-subject differences for each feature in each cohort. EEGET-ALS shows systematically lower between-subject variance proportions, particularly for higher-frequency features. For spec_beta, the between-EEGET-ALS-and-UCL gap is 78 percentage points (15.6% vs 93.7%) and the between-EEGET-ALS-and-HC gap is 68 percentage points (15.6% vs 83.7%) – same feature, same algorithm, different variance structures across cohorts.

### 3.6 Disease-specific instability is rare and restricted to non-discriminating features

Two features, sample entropy and modulation index alpha-gamma, showed high ICC in healthy adults (0.77 and 0.70, respectively) but near-zero or negative ICC in both ALS cohorts. However, neither feature passed criterion 1 (sample entropy: d = 0.11; mi_alpha_gamma: d = 0.13). Even when genuine ALS-specific instability is observed, it occurs in features that do not discriminate ALS from healthy controls (see the sampen and mi_alpha_gamma rows in (Table 2)).

### 3.7 Phase-amplitude coupling features show cross-cohort sign reversal

Four features passed criterion 1 but failed criterion 3: two phase-amplitude coupling (PAC) measures, mvl_alpha_gamma and mvl_theta_gamma; Hjorth mobility; and C3-C4 beta coherence (Table 2). The delta-band PAC feature mvl_delta_gamma showed the same pattern before it was excluded from the primary panel (Section 2.4). The two retained PAC features were reduced below healthy control levels in EEGET-ALS but elevated several standard deviations above healthy control levels in the UCL cohort. Such large opposite-direction deviations between cohorts in the same disease group suggest strong protocol sensitivity, which limits their portability for cross-site clinical deployment.

### 3.8 Threshold sensitivity

Threshold sensitivity analysis (Fig. 6) shows the number of features passing each criterion as a function of ICC threshold (0.3 to 0.8). Features passing criterion 2 declined smoothly from 10 at ICC > 0.4 to 0 at ICC > 0.8. The number passing all three criteria reached 3 at ICC > 0.5-0.6 and dropped to 0 by ICC > 0.675, consistent with sample-size limits on ICC estimation. Several features have ICC point estimates of 0.6-0.65 with bootstrap CIs spanning that range. The smooth curve shape indicates that the primary results are not artefacts of an arbitrary threshold choice; they are stable across a range of moderate thresholds (0.5-0.6).

**Figure 6.**
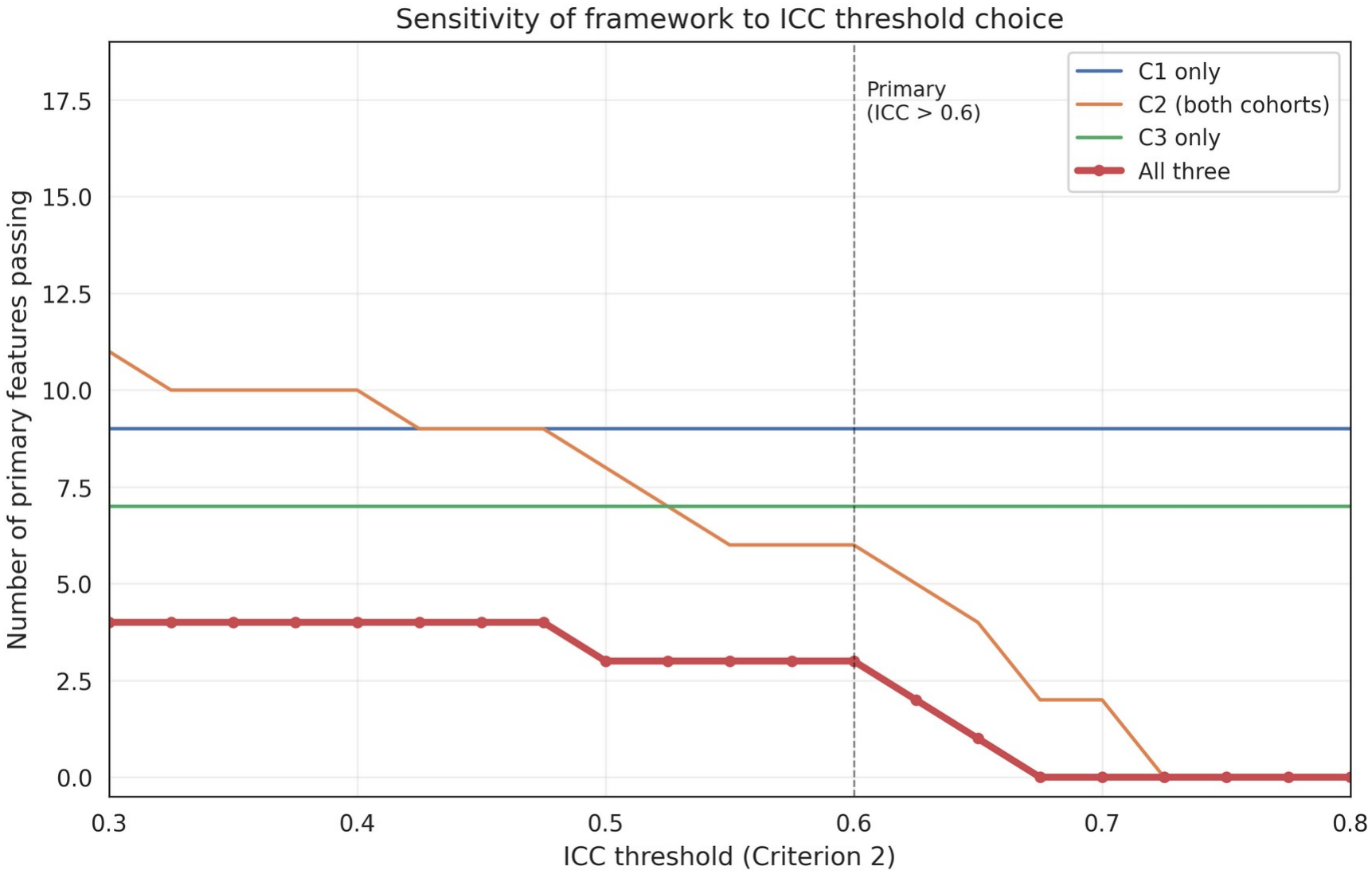
Threshold sensitivity. Number of features passing each criterion as a function of the ICC threshold. The number of features passing Criterion 2 (reliability in both ALS cohorts) declines smoothly with threshold; the number passing all three criteria reaches 3 at moderate threshold (ICC > 0.6) and 0 at strict threshold (ICC > 0.7). The fall to zero at the strict threshold is consistent with sample-size limits on ICC estimation rather than with feature invalidity.

## 4. Discussion

Before summarising the empirical findings, we emphasise the intended scope of the framework. The three criteria (|d| > 0.5, ICC(3,1) > 0.6 in both ALS cohorts, cross-cohort sign concordance) are decision rules for a screening procedure, not claims about biological truth or definitive validation. Their role is to identify a small number of features that jointly satisfy discrimination, reliability, and cross-cohort consistency well enough to justify prospective testing in larger age- and sex-matched ALS cohorts. A feature that fails one or more criteria in this dataset is not necessarily invalid as an ALS biomarker; conversely, a feature that passes all three is a candidate for validation, not a validated biomarker. This screening-vs-validation distinction is central to the interpretation of the results below.

### Summary of findings

Across three independent cohorts spanning two continents and three recording protocols, we applied a pre-registered three-criterion screening framework adapted from the FDA-NIH BEST structure to evaluate 18 EEG-derived candidate biomarkers for ALS [3, 4]. Given the limited sample size (14 ALS participants across two cohorts), this study was explicitly exploratory and underpowered for definitive biomarker validation; the uncertainty around the number of features surviving all three criteria is substantial, as reflected in bootstrap confidence intervals that include zero.

Within these constraints, three main empirical patterns emerged. First, gamma power, theta power, and spectral entropy emerged as framework-positive by meeting all three pre-specified criteria. A mechanistic interpretation consistent with this pattern is altered cortical excitability; increased gamma activity and reduced alpha activity have both been proposed as electrophysiological correlates of excitatory-inhibitory imbalance in ALS [4, 32], although our cross-sectional design does not directly test this hypothesis.

Second, resampling analyses indicated that feature inclusion in the selected set was sensitive to sampling variability. Gamma power, theta power, and spectral entropy were the most consistently recovered features across bootstrap iterations. However, the instability of inclusion across resamples highlights the dependence of candidate selection on sample composition in small-n multi-cohort EEG studies [3].

Third, substantial differences in feature reliability across recording protocols were observed. Several features had higher within-cohort reliability in clinic-based recordings than in home-based recordings, consistent with prior evidence that EEG metrics are sensitive to acquisition context, signal-to-noise conditions, and preprocessing pipelines [11].

### Implications for ALS EEG biomarker development

A central implication is that EEG-derived biomarkers in ALS cannot be generalised across recording contexts without explicit evaluation. Features that appear stable in clinic-based datasets may not retain equivalent properties in home-based recordings. The high cross-cohort reliability of alpha-band measures is notable given that reduced alpha power has been among the most frequently reported EEG findings in ALS [4, 7, 9]; reliability alone, however, was not sufficient for these features to pass the full framework in this dataset.

Pooling EEG data across studies with differing acquisition protocols without explicit modelling of protocol effects may introduce systematic confounding. Future work should therefore incorporate the acquisition protocol as an explicit factor in analytical models rather than treating it as a nuisance variable, report preprocessing pipelines in machine-readable form to enable exact reproduction, evaluate discriminative performance and within-subject reliability jointly rather than in isolation, and, where possible, use multi-cohort designs with harmonised or explicitly contrasted recording conditions. Biomarker evaluation should be aligned with the intended deployment context: features validated under clinic-based acquisition cannot be assumed to generalise to home-based monitoring without additional validation in the intended setting.

### Clinical relevance

Current ALS outcome measures include functional scales such as the ALSFRS-R and blood-based biomarkers such as neurofilament light chain [3]. EEG-based biomarkers could in principle complement these by providing direct cortical physiology measures with higher-frequency longitudinal sampling. The portability and low cost of EEG make it particularly attractive for decentralised monitoring and longitudinal biomarker development in ALS, provided that protocol-related variance is explicitly characterised [4].

### Demographic sensitivity analyses

The EEGET-ALS cohort exhibited a mean age difference of approximately 16 years between ALS and HC groups and a sex composition imbalance (ALS 3M/3F; HC 170M/0F). Post-hoc demographic sensitivity analyses (Table S5) evaluated whether the three selected features survived demographic adjustment under five specifications: the pre-registered original, age-adjusted regression, HC restricted to ALS age range, male-only ALS versus all-male HC, and the most conservative combined specification. Gamma power passed criterion 1 in all five specifications (d ranging from +0.65 to +1.26). Spectral entropy passed in all five specifications with strikingly stable effect sizes (d = +0.71 to +0.84). Theta power passed in four of five specifications but failed in the age-restricted HC condition (d = -0.27); this pattern is consistent with, but not sufficient to establish, a disease-associated component alongside a partial age confound, particularly given the small subgroup sizes involved (only three male ALS participants in the male-only specification).

### Limitations

This study was limited by the small ALS sample size (n = 14 across two cohorts), demographic mismatch between groups in EEGET-ALS, and incomplete longitudinal sampling. Clinical metadata (ALSFRS-R scores, disease duration) were not available for all cohorts, preventing systematic evaluation of symptom-biomarker relationships. Features were aggregated to the recording level by the mean rather than the median specified in the pre-registration, a disclosed deviation (see Methods 2.3); whether the framework-positive set is sensitive to this choice has not been tested and is left as an open question for future replication rather than a planned pre-submission re-run. Future studies should use larger multi-site cohorts (n >= 50), age- and sex-matched controls, harmonised acquisition protocols, and longitudinal follow-up. These results should be interpreted as exploratory screening outputs rather than definitive biomarker validation.

## 5. Conclusion

This study presents a pre-registered, multi-cohort EEG biomarker screening framework for ALS. Within the constraints of a small and heterogeneous sample, gamma power, theta power, and spectral entropy emerged as relatively stable candidates across analytic criteria and recording contexts. The principal methodological finding is that cohort-associated variability – plausibly including a substantial acquisition-protocol contribution, though protocol is confounded with cohort, equipment, setting, and preprocessing in this design and cannot be isolated from them – was large relative to the apparent disease-related signal, making explicit protocol characterisation important in multi-cohort biomarker research. Rather than establishing validated biomarkers, this study provides a structured screening approach for identifying EEG features that warrant further evaluation in larger, harmonised, and longitudinal ALS studies.

## Author contributions

Belay Gebregergis: Conceptualisation, Methodology, Software, Formal analysis, Data curation, Writing - original draft, Visualisation. Haben Girmay Yhdego: Software, Writing - review and editing. Tewolde Teklu: Conceptualisation, Supervision, Writing - review and editing.

## Acknowledgments

We thank the original investigators of the EEGET-ALS, UCL/Penn State, and HC TestRetest datasets for making their data publicly available; this cross-cohort screening analysis would not have been possible without them.

## Ethics statement

This study was a secondary analysis of three publicly available, fully de-identified EEG datasets: EEGET-ALS [12], UCL/Penn State [6, 13], and HC TestRetest [11]. Ethics approval and informed consent for each dataset were obtained by the respective original investigators and documented in primary publications. No new data were collected from human participants and no identifying information was accessed at any stage.

## Data and code availability

The EEGET-ALS data are publicly available via Figshare [12] (doi:10.6084/m9.figshare.c.6910027). UCL/Penn State data are publicly available on Figshare [13] (doi:10.5522/04/28156016). HC TestRetest data are publicly available on OpenNeuro (ds004148) [11]. All analysis code is available at https://github.com/BelayTG/eeg-ml-eeget-als. The pre-registered analysis framework is available at OSF (doi:10.17605/OSF.IO/4K32S).

