## Supplementary Material for "Cross-cohort validation of EEG biomarkers using a pre-registered three-criterion framework: a demonstration in amyotrophic lateral sclerosis"

Supplementary Materials

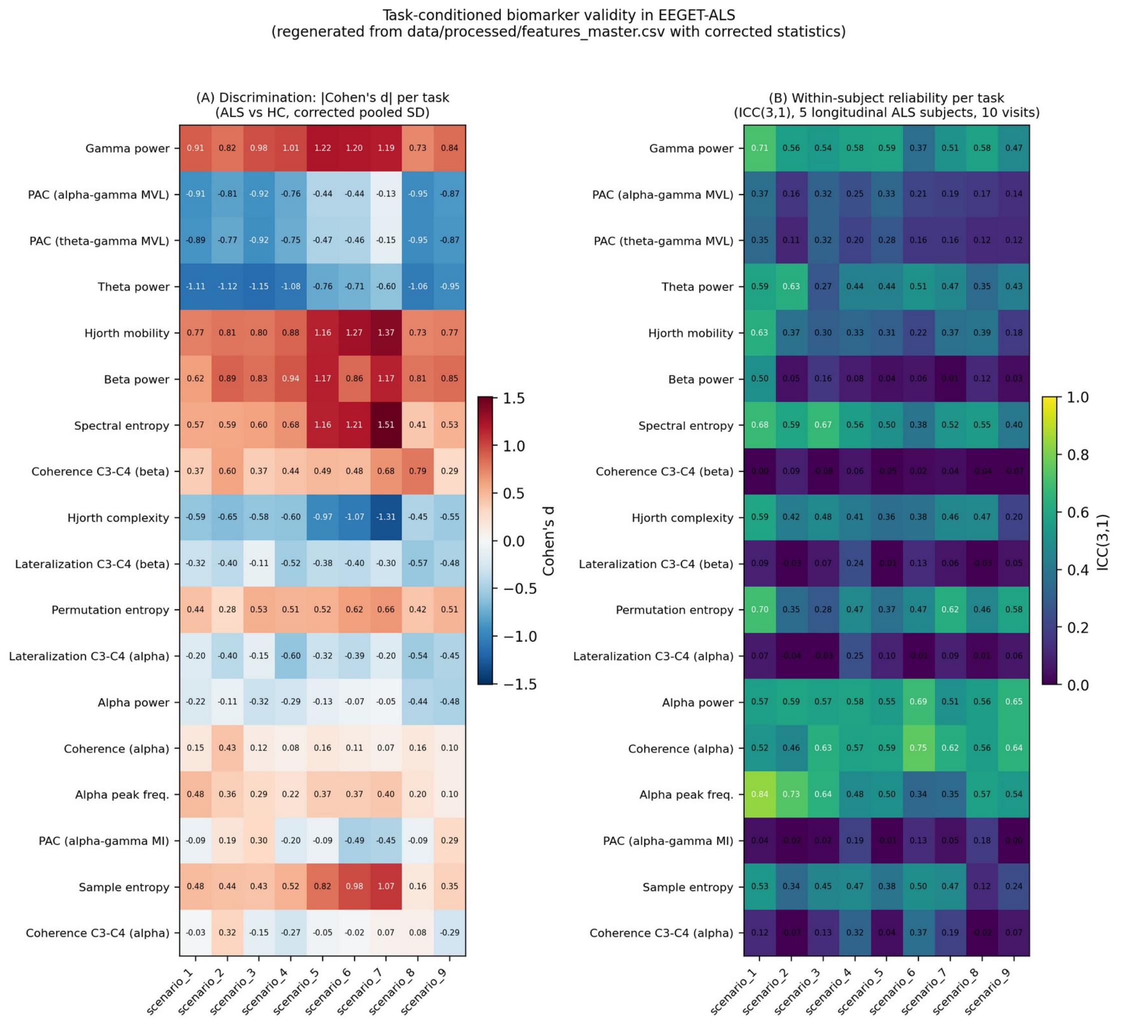


*Figure S1*

Figure S1: Task-conditioned biomarker validity in EEGET-ALS, computed with `scripts/generate_figure_s1.py` directly from `data/processed/features_master.csv` using the same corrected statistics (sample-size-weighted pooled-SD Cohen's d; two-way mixed-effects, absolute-agreement ICC(3,1)) used throughout this manuscript. This replaces an earlier version of this figure that had no generating script in the tracked repository and whose Cohen's d values could not be confirmed to use the corrected formula; that earlier version is superseded. (A) Discrimination panel. Per-feature Cohen's d (ALS vs. HC), computed separately within each of the nine EEGET-ALS task scenarios, for all 18 primary features. (B) Within-subject reliability panel. Per-feature ICC(3,1), computed across the 10 longitudinal visits of the five longitudinal EEGET-ALS ALS subjects, estimated independently within each of the nine task scenarios. Together, panels A and B characterize whether candidate biomarkers retain validity uniformly across task conditions or are conditionally sensitive to the specific acquisition scenario, a consideration for deployment in heterogeneous monitoring contexts. Task scenarios are labeled `scenario_1`-`scenario_9`, matching the `task` column of `features_master.csv` verbatim; the manuscript's description of these as four limb motor-imagery tasks, three bulbar/cervical movement tasks, and two augmentative communication intents (Methods 2.2) reflects the study design, but the specific scenario-number-to-task-label correspondence is not recorded anywhere in the tracked repository and so is not asserted here. The underlying values are in `results/per_task_validity.csv`.

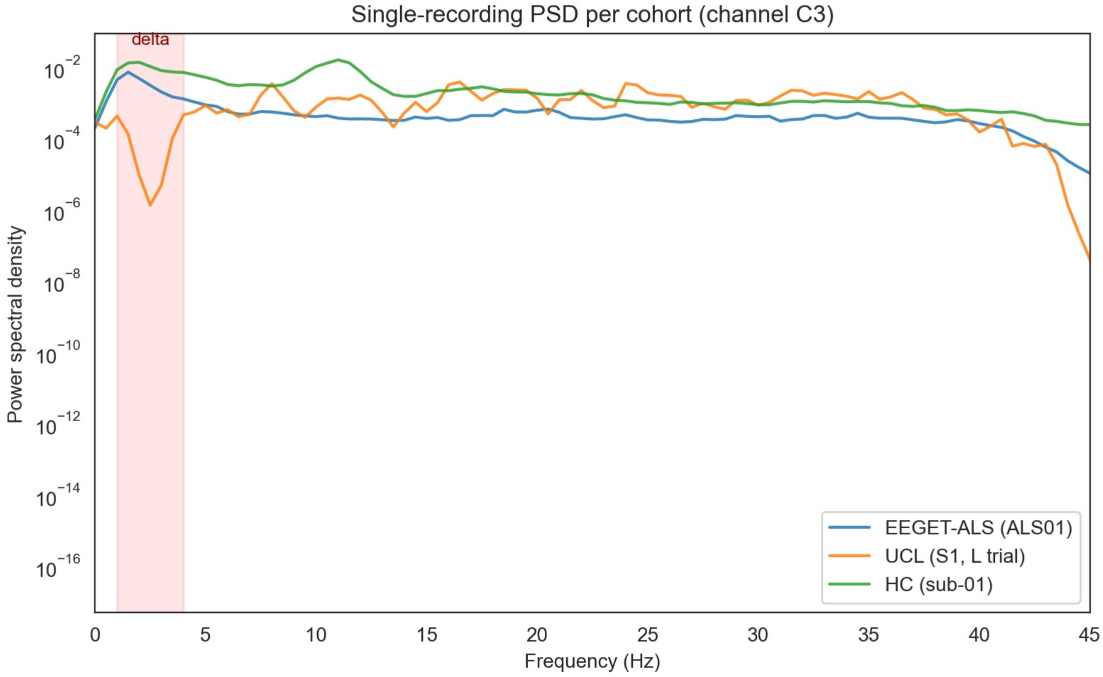


*Figure S2*

Figure S2: Cross-cohort power spectral density diagnostic showing the UCL delta-band notch that motivated excluding two delta-band features from primary analysis.

Table S1. Literature provenance for each of the 18 features in the pre-registered primary panel that entered the cross-cohort analysis. Each feature is annotated with a representative ALS-EEG (or where ALS-specific evidence is sparse, neurodegeneration-EEG) publication. Citations were identified through targeted literature search and cross-validated against the Chmiel & Stępień-Słodkowska 2025 systematic review of resting-state EEG in ALS (J Clin Med 14:545; doi:10.3390/jcm14020545), with primary sources cited directly where the review identified them. These citations are not exhaustive and not necessarily the original or definitive sources for each feature; they indicate prior examination of the feature in ALS or related neurodegenerative-disease EEG biomarker research. Confidence column reflects strength of direct ALS-specific evidence: High = direct ALS study reporting the feature with clear between-group effects; Moderate = ALS study reporting a related variant or broader claim than the specific measurement used here; Low = ALS-specific evidence sparse, feature is panel-motivated by general EEG biomarker literature; Low (ALS); High or Moderate (general) = feature is a well-established EEG biomarker in broader neurodegeneration literature but ALS-specific direct evidence is sparse. Excluded delta-band features are documented in Table S2; exploratory channel-pair variants are documented in Table S4.

| Feature | Family | Claim / Role in ALS-EEG Literature | Primary Source(s) | Journal(s) | Confidence |
|---|---|---|---|---|---|
| alpha_peak_freq | Spectral | Alpha peak frequency slows markedly in late-stage ALS patients. APF values of 7 Hz and 3.8 Hz have been recorded in CLIS patients, well below the normal 8-13 Hz range (Hohmann 2018). Pronounced frequency slowing with reduced or absent alpha activity is also reported in CLIS (Khalili-Ardali 2021). | Hohmann et al. 2018 (Clin Neurophysiol 129:406-408, doi:10.1016/j.clinph.2017.11.013); Khalili-Ardali et al. 2021 (Clin Neurophysiol 132:1064-1076, doi:10.1016/j.clinph.2021.02.002) | Clin Neurophysiol (both) | High |
| spec_alpha | Spectral | Reduced alpha-band power is the most consistently reported resting-state EEG finding in ALS and the headline result of the Chmiel 2025 review. Alpha reductions have been observed at central electrodes C3, C4, and Cz (Mai 1998); left-hemisphere power is 60-70% lower than in controls (Santhosh 2005); parietal and occipital reductions are reported by Deligani 2020; a central trend reduction by Jayaram 2015; and reduced alpha2/alpha3 source power in LIS patients by Babiloni 2010. | Mai et al. 1998 (Electroencephalogr Clin Neurophysiol 106:383-386, no DOI - pre-digital); Santhosh et al. 2005 (Neurol India 53:99-101, doi:10.4103/0028-3886.15071); Deligani et al. 2020 (IEEE TNSRE, doi:10.1109/TNSRE.2020.3031495); Jayaram et al. 2015 (IEEE EMBC); Babiloni et al. 2010 (Clin Neurophysiol 121:1816-1824, doi:10.1016/j.clinph.2010.04.027) | Multiple (Electroencephalogr Clin Neurophysiol; Neurol India; IEEE TNSRE; IEEE EMBC; Clin Neurophysiol) | High |
| spec_beta | Spectral | Beta-band power changes in ALS track disease severity and motor symptom progression. A positive | Notturno et al. 2023 (Amyotroph Lateral Scler Frontotemporal | Multiple (ALS-FTD; Eur J Neurol; Brain; | High |

| Feature | Family | Claim / Role in ALS-EEG Literature | Primary Source(s) | Journal(s) | Confidence |
|---|---|---|---|---|---|
| | | correlation with progression scores has been found in motor and frontal regions (Notturno 2023). Altered sensorimotor beta power correlates with lower motor neuron impairment and faster progression (Dukic 2024). Increased beta power in the frontotemporal network characterises one distinct ALS subphenotype (Dukic 2022), and reduced beta-band synchrony is seen in the ALSncbi subgroup (Metzger 2024). | Degener 24:295-303, doi:10.1080/21678421.2022.); Dukic et al. 2024 (Eur J Neurol 31:e16201, doi:10.1111/ene.16201); Dukic et al. 2022 (Brain 145:621-631, doi:10.1093/brain/awab322); Metzger et al. 2024 (Brain Topogr 38:3, doi:10.1007/s10548-024-01080-0) | Brain Topogr) | |
| spec_ gamma | Spectral | Gamma-band changes in ALS are subphenotype-specific and regionally variable. Four EEG subphenotypes show distinct gamma signatures including frontotemporal gamma reduction and frontoparietal gamma comodulation (Dukic 2022). Enhanced global gamma power in cortical regions is reported by Jayaram 2015; region-specific gamma increases over frontoparietal areas by Nasseroleslami 2019; and elevated gamma in frontal, parietal, and occipital regions by Iyer 2015. Longitudinal gamma increases in | Dukic et al. 2022 (Brain 145:621-631, doi:10.1093/brain/awab322); Jayaram et al. 2015 (IEEE EMBC); Nasseroleslami et al. 2019 (Cereb Cortex 29:27-41, doi:10.1093/cercor/bhx301); Iyer et al. 2015 (PLOS ONE 10:e0128682, doi:10.1371/journal.pone.0128682); Metzger et al. 2024 (Brain Topogr 38:3, doi:10.1007/s10548-024-01080-0) | Multiple (Brain; IEEE EMBC; Cereb Cortex; PLOS ONE; Brain Topogr) | High |

| Feature | Family | Claim / Role in ALS-EEG Literature | Primary Source(s) | Journal(s) | Confidence |
|---|---|---|---|---|---|
| | | frontal and temporal lobes, with an ALSbi-specific temporal lobe rise, are described by Metzger 2024. | | | |
| spec_theta | Spectral | Theta-band power changes in ALS are directionally inconsistent across studies and appear region-dependent. Reductions over bilateral motor areas are reported by Nasseroleslami 2019, and in frontal, central, and temporal regions by Deligani 2020. Conversely, elevated theta in central, frontal, and parietal regions is described by Iyer 2015, while occipital and temporal reductions appear in Dukic 2019. The direction of change depends on the region of interest and the analysis approach used. | Nasseroleslami et al. 2019 (Cereb Cortex 29:27-41, doi:10.1093/cercor/bhx301); Deligani et al. 2020 (IEEE TNSRE, doi:10.1109/TNSRE.2020.3032536); Iyer et al. 2015 (PLOS ONE 10:e0128682, doi:10.1371/journal.pone.0128682); Dukic et al. 2019 (Hum Brain Mapp, doi:10.1002/hbm.24740) | Multiple (Cereb Cortex; IEEE TNSRE; PLOS ONE; Hum Brain Mapp) | High |
| spectral_entropy | Spectral | Spectral entropy is a general EEG complexity and irregularity measure with established relevance to neurodegeneration and aging. Its inclusion here is motivated by its role as an entropy-class measure in the broader neurodegenerative disease literature. | Cacciotti, Pappalettera, Miraglia, Rossini & Vecchio 2024 (GeroScience 46:5537-5557, doi:10.1007/s11357-024-01185-1) | GeroScience (review) | Low (ALS-specific); High (general EEG complexity) |
| mi_alpha_ | PAC | Modulation index quantification of alpha- | Benetton et al. 2025 (Brain | Brain Communic | Moderate |

| Feature | Family | Claim / Role in ALS-EEG Literature | Primary Source(s) | Journal(s) | Confidence |
|---|---|---|---|---|---|
| gamma | | gamma phase-amplitude coupling is an exploratory ALS candidate with emerging but limited direct evidence. | Communications 7:fcaf192, doi:10.1093/braincomms/fcaf192) | ations | |
| mvl_alpha_gamma | PAC | Alpha-gamma phase-amplitude coupling, quantified using the mean vector length metric, is altered in the ALS motor and sensorimotor cortex. This supports inclusion of MVL-based alpha-gamma coupling as a candidate feature. | Benetton et al. 2025 (Brain Communications 7:fcaf192, doi:10.1093/braincomms/fcaf192) | Brain Communications | High |
| mvl_theta_gamma | PAC | Theta-gamma phase-amplitude coupling is reduced in the dominant motor cortex in ALS and has been linked to cortical hyperexcitability and noradrenaline deficiency. | Scekic-Zahirovic et al. 2024 (Sci Transl Med, doi:10.1126/scitranslmed.adg3665); Benetton et al. 2025 (Brain Communications 7:fcaf192, doi:10.1093/braincomms/fcaf192) | Sci Transl Med; Brain Communications | High |
| hjorth_complexity | Complexity | Hjorth complexity is a normalised slope descriptor that estimates EEG spectral bandwidth by comparing how closely a signal resembles a pure sine wave. It is a well-established time-domain EEG feature, but no strong ALS-specific validation has been identified. | Hjorth 1970 (Electroencephalogr Clin Neurophysiol 29:306-310, doi:10.1016/0013-4694(70)90143-4) | Electroencephalogr Clin Neurophysiol | Low (ALS-specific) |
| hjorth_ | Complex | Hjorth mobility | Hjorth 1970 | Electroenc | Low (ALS- |

| Feature | Family | Claim / Role in ALS-EEG Literature | Primary Source(s) | Journal(s) | Confidence |
|---|---|---|---|---|---|
| mobilit y | ity | estimates mean frequency as the proportion of standard deviation of the power spectrum. It is a well-established time-domain EEG feature, but no strong ALS-specific validation has been identified. | (Electroencephalogr Clin Neurophysiol 29:306-310, doi:10.1016/0013-4694(70)90143-4) | ephalogr Clin Neurophysiol | specific) |
| peen | Complexity | Permutation entropy is a general EEG complexity and irregularity biomarker with relevance to aging and neurodegeneration. ALS-specific direct evidence has not been identified. Its inclusion is motivated by the broader entropy-based EEG complexity literature in neurodegenerative disease. | Cacciotti, Pappalettera, Miraglia, Rossini & Vecchio 2024 (GeroScience 46:5537-5557, doi:10.1007/s11357-024-01185-1) | GeroScience (review) | Low (ALS-specific); Moderate (general) |
| sampen | Complexity | Sample entropy is a general EEG complexity and irregularity biomarker. ALS-specific direct evidence has not been identified. Its inclusion is motivated by its established role as a core entropy measure in the EEG complexity literature for neurodegenerative disease. | Cacciotti, Pappalettera, Miraglia, Rossini & Vecchio 2024 (GeroScience 46:5537-5557, doi:10.1007/s11357-024-01185-1) | GeroScience (review) | Low (ALS-specific); Moderate (general) |
| coh_ alpha | Connectivity | Whole-scalp alpha-band functional connectivity is broadly altered in ALS. Iyer 2015 reports increased alpha cross- | Iyer et al. 2015 (PLOS ONE 10:e0128682, doi:10.1371/journal.pone.012868 | Multiple (PLOS ONE; Brain) | High |

| Feature | Family | Claim / Role in ALS-EEG Literature | Primary Source(s) | Journal(s) | Confidence |
|---|---|---|---|---|---|
| | | spectral density across the scalp and increased alpha-band clustering coefficient in all scalp regions. Dukic 2022 identifies altered alpha-band synchrony in the somatomotor network as a marker of one distinct ALS subphenotype. | 2); Dukic et al. 2022 (Brain 145:621-631, doi:10.1093/brain/awab322) | | |
| coh_alpha_C3_C4 | Connectivity | Alpha-band coherence between sensorimotor regions C3 and C4 is altered in ALS. Increased alpha-band synchrony in the somatomotor network characterises one distinct ALS subphenotype (Dukic 2022). Characteristic increases in EEG connectivity that correlate with structural MRI changes in ALS are reported by Nasseroleslami 2019. Broadly increased alpha cross-spectral density across the scalp, including sensorimotor regions, is shown by Iyer 2015. | Dukic et al. 2022 (Brain 145:621-631, doi:10.1093/brain/awab322); Nasseroleslami et al. 2019 (Cereb Cortex 29:27-41, doi:10.1093/cercor/bhx301); Iyer et al. 2015 (PLOS ONE 10:e0128682, doi:10.1371/journal.pone.0128682) | Multiple (Brain; Cereb Cortex; PLOS ONE) | High |
| coh_beta_C3_C4 | Connectivity | Beta-band connectivity between sensorimotor regions C3 and C4 is altered in ALS. Increased beta-band spectral power in the frontotemporal network characterises one distinct ALS subphenotype (Dukic 2022). Resting-state EEG | Dukic et al. 2022 (Brain 145:621-631, doi:10.1093/brain/awab322); Iyer et al. 2015 (PLOS ONE 10:e0128682, doi:10.1371/journal.pone.0128682); Dukic et | Multiple (Brain; PLOS ONE; Eur J Neurol) | Moderate |

| Feature | Family | Claim / Role in ALS-EEG Literature | Primary Source(s) | Journal(s) | Confidence |
|---|---|---|---|---|---|
| | | functional connectivity changes across frequency bands as a potential ALS biomarker are documented by Iyer 2015. Altered sensorimotor beta-band power correlating with motor symptom severity is reported by Dukic 2024. | al. 2024 (Eur J Neurol 31:e16201, doi:10.1111/ene.16201) | | |
| lat_alpha_C3_C4 | Lateralization | Alpha power is lateralised between motor cortex electrodes C3 and C4 in ALS. Left-hemisphere alpha power is 60-70% lower in ALS patients than in controls, with the reduction more pronounced on the left than the right (Santhosh 2005). Reduced relative alpha power over the motor cortex is also reported by Mai 1998, though C4 specificity should be confirmed against the original paper. Geronimo 2016 provides motivation for the C3/C4 panel from BCI and motor lateralization analyses. | Santhosh et al. 2005 (Neurol India 53:99-101, doi:10.4103/0028-3886.15071); Mai et al. 1998 (Electroencephalogr Clin Neurophysiol 106:383-386, no DOI - pre-digital); Geronimo et al. 2016 (J Neural Eng 13:026002, doi:10.1088/1741-2560/13/2/026002) | Multiple (Neurol India; Electroencephalogr Clin Neurophysiol; J Neural Eng) | Moderate |
| lat_beta_C3_C4 | Lateralization | Beta-band lateralization between C3 and C4 is included in the panel on the basis of BCI and motor lateralization analyses in ALS patients (Geronimo 2016), together with region-specific beta-band changes in sensorimotor | Geronimo et al. 2016 (J Neural Eng 13:026002, doi:10.1088/1741-2560/13/2/026002); Dukic et al. 2022 (Brain 145:621-631, | Multiple (J Neural Eng; Brain; Eur J Neurol) | Moderate |

| Feature | Family | Claim / Role in ALS-EEG Literature | Primary Source(s) | Journal(s) | Confidence |
|---|---|---|---|---|---|
| | | areas reported across ALS studies (Dukic 2022, 2024). Direct evidence for C3/C4 beta lateralization as a standalone ALS biomarker remains limited. | doi:10.1093/brain/awab322); Dukic et al. 2024 (Eur J Neurol 31:e16201, doi:10.1111/ene.16201) | | |

Notes for Table S1: Of the 18 primary features that entered analysis, the confidence breakdown is: High (10): direct ALS-EEG studies reporting the feature with clear between-group effects; Moderate (3): ALS evidence for a related variant or broader claim than the specific measurement used here; Low (ALS); High or Moderate (general) (3): well-established EEG biomarkers in broader neurodegeneration literature with sparse direct ALS evidence (spectral_entropy, sampen, peen); Low (2): Hjorth mobility and Hjorth complexity, retained as general EEG features without strong ALS-specific direct evidence. The three selected features (framework-positive) span confidence tiers: gamma power (High), theta power (High), and spectral entropy (Low ALS; High general). This table is provided for transparency about the literature anchoring of each feature; it does not replace or modify the pre-registered framework.

Table S2. Excluded delta-band features. The two pre-registered delta-band features were excluded from the primary 18-feature panel after pilot inspection revealed an order-of-magnitude reduction in UCL relative delta-band power consistent with undocumented preprocessing in this band. Any apparent ALS-vs-HC discrimination on these features is uninterpretable due to the cross-cohort spectral confound.

| Feature | Family | Cohen's d [95% CI] | ICC EG [95% CI] | ICC UCL [95% CI] | ICC HC [95% CI] |
|---|---|---|---|---|---|
| spec_delta | Spectral | −0.47 [-1.29, 0.25] | 0.58 [-0.11, 0.75] | 0.79 [0.29, 0.89] | 0.50 [0.32, 0.65] |
| mvl_delta_gamma | PAC | −1.08 [-1.60, -0.65] | 0.25 [-0.09, 0.51] | 0.59 [0.11, 0.83] | 0.43 [0.26, 0.55] |

Table S3. Cross-cohort z-score sensitivity check on Criterion 3. For each feature, z-scores represent the ALS mean's deviation from the EEGET-ALS HC distribution. Criterion 3 'original' uses same-sign AND |z| > 0.1 for both cohorts. Criterion 3 'no floor' uses only the sign-concordance requirement. Yellow-shaded rows indicate features whose C3 status changes when the |z| > 0.1 floor is removed.

| Feature | Panel | EG z to HC | UCL z to HC | Same sign? | C3 with \|z\| >0.1 | C3 no floor |
|---|---|---|---|---|---|---|
| mvl_theta_gamma | Primary | −0.87 | +2.71 | - | - | - |

| Feature | Panel | EG z to HC | UCL z to HC | Same sign? | C3 with \|z\| >0.1 | C3 no floor |
|---|---|---|---|---|---|---|
| mvl_alpha_gamma | Primary | −0.88 | +1.79 | - | - | - |
| spec_gamma | Primary | +0.95 | +2.80 | √ | √ | √ |
| coh_beta_C3_C4 | Primary | +0.71 | +0.07 | √ | - | √ |
| spec_beta | Primary | +0.79 | +5.30 | √ | √ | √ |
| hjorth_mobility | Primary | +0.80 | −2.00 | - | - | - |
| spec_theta | Primary | −0.87 | −0.60 | √ | √ | √ |
| spectral_entropy | Primary | +0.75 | +0.38 | √ | √ | √ |
| hjorth_complexity | Primary | −0.66 | −3.27 | √ | √ | √ |
| peen | Primary | +0.41 | −11.12 | - | - | - |
| lat_beta_C3_C4 | Primary | −0.41 | −0.05 | √ | - | √ |
| lat_alpha_C3_C4 | Primary | −0.31 | +0.22 | - | - | - |
| spec_alpha | Primary | −0.28 | +0.47 | - | - | - |
| coh_alpha | Primary | +0.19 | −1.75 | - | - | - |
| mi_alpha_gamma | Primary | +0.13 | +0.54 | √ | √ | √ |
| alpha_peak_freq | Primary | +0.13 | +0.59 | √ | √ | √ |
| coh_alpha_C3_C4 | Primary | −0.07 | −0.41 | √ | - | √ |
| sampen | Primary | +0.11 | −1.81 | - | - | - |
| mvl_delta_gamma | Exploratory | −0.84 | +3.03 | - | - | - |
| coh_beta | Exploratory | +0.76 | −1.27 | - | - | - |
| spec_delta | Exploratory | −0.50 | −5.54 | √ | √ | √ |
| lat_beta_Fp1_Fp2 | Exploratory | +0.27 | +0.21 | √ | √ | √ |
| mi_delta_gamma | Exploratory | +0.19 | +0.55 | √ | √ | √ |
| lat_alpha_Fp1_Fp2 | Exploratory | +0.21 | +0.02 | √ | - | √ |
| lat_beta_F3_ | Exploratory | −0.17 | −0.08 | √ | - | √ |

| Feature | Panel | EG z to HC | UCL z to HC | Same sign? | C3 with \|z\| >0.1 | C3 no floor |
|---|---|---|---|---|---|---|
| F4 | | | | | | |
| mi_theta_gamma | Exploratory | +0.11 | +0.55 | √ | √ | √ |
| lat_alpha_F3_F4 | Exploratory | −0.10 | −0.05 | √ | - | √ |

Notes for Table S3: Removing the |z| > 0.1 floor adds 6 features to the C3-passing set, but none of these features pass Criterion 2 (within-subject reliability). The three priority candidates remain unchanged under this sensitivity check.

Table S4. Full per-feature scorecard for all 27 features common to all three cohorts. Primary panel = 18 pre-registered features; Exploratory panel (gray-shaded, italicized) = 9 channel-pair variants requiring EEGET-ALS-specific electrodes, retained for transparency. Cohen's d computed on subject-averaged EEGET-ALS data (ALS vs HC). ICC EG = ICC(3,1) in EEGET-ALS visit-level data; ICC UCL = ICC(3,1) in UCL session-level data. C1 = passes discrimination (|d| > 0.5). C2 = passes reliability in both ALS cohorts. C3 = passes cross-cohort sign concordance with |z| > 0.1 floor.

| Feature | Family | Panel | Cohen's d [95% CI] | ICC EG [95% CI] | ICC UCL [95% CI] | C1 | C2 | C3 |
|---|---|---|---|---|---|---|---|---|
| spec_gamma | Spectral | Primary | +0.95 [+0.35, +1.80] | 0.67 [0.04, 0.81] | 0.84 [0.60, 0.93] | √ | √ | √ |
| mvl_alpha_gamma | PAC | Primary | -0.89 [-1.29, -0.48] | 0.31 [-0.03, 0.55] | 0.53 [0.04, 0.79] | √ | - | - |
| mvl_theta_gamma | PAC | Primary | -0.88 [-1.26, -0.52] | 0.26 [-0.05, 0.54] | 0.69 [0.17, 0.86] | √ | - | - |
| spec_theta | Spectral | Primary | -0.87 [-1.64, -0.16] | 0.60 [0.07, 0.77] | 0.82 [0.33, 0.92] | √ | √ | √ |
| hjorth_mobility | Complexity | Primary | +0.80 [+0.25, +1.47] | 0.42 [-0.07, 0.72] | 0.82 [0.43, 0.90] | √ | - | - |
| spec_beta | Spectral | Primary | +0.79 [+0.25, +1.39] | 0.09 [-0.10, 0.51] | 0.93 [0.52, 0.96] | √ | - | √ |
| spectral_entropy | Spectral | Primary | +0.75 [+0.06, | 0.64 [0.17, | 0.71 [0.27, | √ | √ | √ |

| Feature | Family | Panel | Cohen’s d [95% CI] | ICC EG [95% CI] | ICC UCL [95% CI] | C 1 | C 2 | C 3 |
|---|---|---|---|---|---|---|---|---|
| | | | +1.56] | 0.82] | 0.80] | | | |
| coh_beta_C3_C4 | Connectivity | Primary | +0.72 [+0.38, +1.34] | 0.03 [-0.09, 0.21] | 0.83 [0.54, 0.89] | √ | - | - |
| hjorth_ complexity | Complexity | Primary | -0.67 [-1.36, -0.08] | 0.49 [-0.04, 0.76] | 0.87 [-0.03, 0.95] | √ | - | √ |
| lat_beta_C3_C4 | Lateralization | Primary | -0.42 [-0.83, +0.00] | 0.21 [-0.06, 0.49] | 0.04 [-0.14, 0.43] | - | - | - |
| peen | Complexity | Primary | +0.41 [+0.00, +0.80] | 0.65 [0.06, 0.73] | 0.55 [0.14, 0.72] | - | - | - |
| lat_alpha_C3_C4 | Lateralization | Primary | -0.31 [-0.73, +0.18] | 0.17 [-0.11, 0.42] | 0.33 [-0.04, 0.67] | - | - | - |
| spec_alpha | Spectral | Primary | -0.29 [-0.74, +0.23] | 0.66 [0.13, 0.81] | 0.88 [0.57, 0.92] | - | √ | - |
| coh_alpha | Connectivity | Primary | +0.19 [-0.42, +1.04] | 0.72 [-0.02, 0.86] | 0.73 [0.36, 0.84] | - | √ | - |
| alpha_peak_freq | Spectral | Primary | +0.13 [-0.56, +0.88] | 0.71 [0.13, 0.76] | 0.84 [0.41, 0.94] | - | √ | √ |
| mi_alpha_gamma | PAC | Primary | +0.13 [-0.15, +4.98] | -0.03 [-0.05, 0.28] | -0.01 [-0.05, 0.00] | - | - | √ |
| sampen | Complexity | Primary | +0.11 [-1.21, +1.18] | 0.51 [0.01, 0.76] | 0.60 [0.29, 0.76] | - | - | - |
| coh_alpha_C3_C4 | Connectivity | Primary | -0.07 [-0.36, +0.29] | 0.27 [-0.04, 0.49] | 0.73 [0.36, 0.84] | - | - | - |
| mvl_delta_ gamma | PAC | Exploratory | -0.85 [-1.23, -0.47] | 0.24 [-0.05, | 0.59 [0.13, | √ | - | - |

| Feature | Family | Panel | Cohen's d [95% CI] | ICC EG [95% CI] | ICC UCL [95% CI] | C1 | C2 | C3 |
|---|---|---|---|---|---|---|---|---|
| | | | | 0.52] | 0.83] | | | |
| coh_beta | Connectivity | Exploratory | +0.77 [+0.19, +1.65] | 0.61 [-0.06, 0.79] | 0.83 [0.54, 0.89] | √ | √ | - |
| spec_delta | Spectral | Exploratory | -0.49 [-1.35, +0.22] | 0.57 [-0.09, 0.76] | 0.79 [0.31, 0.89] | - | - | √ |
| lat_beta_Fp1_Fp2 | Lateralization | Exploratory | +0.28 [-0.26, +0.78] | 0.20 [-0.05, 0.38] | -0.16 [-0.28, 0.14] | - | - | √ |
| lat_alpha_Fp1_Fp2 | Lateralization | Exploratory | +0.21 [-0.32, +0.82] | 0.35 [0.03, 0.47] | 0.47 [-0.24, 0.55] | - | - | - |
| mi_delta_gamma | PAC | Exploratory | +0.19 [-0.08, +2.86] | -0.03 [-0.05, 0.30] | -0.01 [-0.05, 0.01] | - | - | √ |
| lat_beta_F3_F4 | Lateralization | Exploratory | -0.17 [-0.78, +0.53] | 0.36 [-0.02, 0.47] | 0.61 [0.14, 0.82] | - | - | - |
| mi_theta_gamma | PAC | Exploratory | +0.12 [-0.16, +4.56] | -0.03 [-0.05, 0.33] | -0.02 [-0.05, 0.00] | - | - | √ |
| lat_alpha_F3_F4 | Lateralization | Exploratory | -0.10 [-0.58, +0.41] | 0.23 [-0.03, 0.32] | 0.76 [0.21, 0.85] | - | - | - |

Notes for Table S4: Three primary features pass all three criteria (spec_gamma, spec_theta, spectral_entropy), the framework-positive set. Among exploratory features, none pass all three criteria.

Table S5. Demographic sensitivity analysis on the pre-registered Criterion 1 (subject-level cross-sectional Cohen's d, ALS vs HC, in EEGET-ALS). Five specifications: (i) d original: pre-registered: 6 ALS vs 170 HC, no demographic adjustment. (ii) d age-adj.: partial standardized group coefficient from OLS regression (feature ~ group + age), using all subjects. (iii) d age-restr. HC: Cohen's d after restricting HC to the ALS age range (37–77; n_HC = 52). (iv) d male-only: Cohen's d after restricting ALS to males only (n_ALS = 3) vs the all-male HC. (v) d combined: most conservative specification: male-only ALS (n = 3) vs age-restricted male HC (n = 52). Robust? column: √√ = passes |d| > 0.5 in all 5 specifications; √ = passes in 4 of 5; ◐ = passes in

2–3 of 5; - = passes in 0–1. Priority candidates (gamma power, spectral entropy, theta power) shown at top of table with colored backgrounds (green = robust across all specifications, yellow = partial).

| Feature | Panel | d original | d age-adj. | d age-restr. HC | d male-only | d combined | Robust? |
|---|---|---|---|---|---|---|---|
| spec_gamma | Primary | +0.95 | +0.86 | +0.65 | +1.26 | +0.95 | √√ |
| spec_theta | Primary | −0.87 | −0.64 | −0.27 | −1.37 | −0.88 | √ |
| spectral_entropy | Primary | +0.75 | +0.84 | +0.74 | +0.73 | +0.71 | √√ |
| mvl_alpha_gamma | Primary | −0.89 | −0.71 | −0.52 | −0.71 | −0.32 | √ |
| mvl_theta_gamma | Primary | −0.88 | −0.71 | −0.53 | −0.68 | −0.30 | √ |
| hjorth_mobility | Primary | +0.80 | +0.72 | +0.46 | +1.25 | +0.85 | √ |
| spec_beta | Primary | +0.79 | +0.65 | +0.32 | +1.22 | +0.68 | √ |
| coh_beta_C3_C4 | Primary | +0.72 | +0.57 | +0.23 | +0.97 | +0.41 | ◐ |
| hjorth_complexity | Primary | −0.67 | −0.84 | −0.72 | −0.88 | −0.87 | √√ |
| lat_beta_C3_C4 | Primary | −0.42 | −0.35 | −0.38 | −0.80 | −0.76 | ◐ |
| peen | Primary | +0.41 | +0.15 | −0.40 | +0.68 | +0.07 | - |
| lat_alpha_C3_C4 | Primary | −0.31 | −0.15 | −0.02 | −0.81 | −0.57 | ◐ |
| spec_alpha | Primary | −0.29 | +0.17 | +1.22 | −0.22 | +1.33 | ◐ |
| coh_alpha | Primary | +0.19 | +0.46 | +0.63 | +0.33 | +0.75 | ◐ |
| alpha_peak_freq | Primary | +0.13 | +0.46 | +0.83 | +0.16 | +0.86 | ◐ |
| mi_alpha_gamma | Primary | +0.13 | +0.18 | +2.16 | +0.11 | +2.66 | ◐ |
| sampen | Primary | +0.11 | +0.31 | +0.37 | +0.67 | +0.82 | ◐ |
| coh_alpha_C3_C4 | Primary | −0.07 | +0.03 | +0.10 | +0.13 | +0.27 | - |

Notes for Table S5: These analyses are post-hoc and not part of the pre-registered framework; they are reported as supplementary sensitivity checks. Gamma power and spectral entropy pass Criterion 1 (|d| > 0.5) in all five specifications; their ALS-associated effect is robust to age and sex adjustment. Theta power passes Criterion 1 in four of five specifications but fails in the age-restricted HC condition (d = −0.27), indicating a partial age confound; the male-only ALS analysis yielded the largest theta effect (d = −1.37), which is consistent with, but given the severe underpowering of that specification (n = 3 ALS) does not by itself establish, a disease-associated component independent of age. Two features that did not pass the pre-registered Criterion 1 (alpha peak frequency, modulation index alpha-gamma) show substantially larger effects under age-restricted HC analysis (+0.83 and +2.16, respectively), indicating that demographic mismatch can suppress as well as inflate effects. The male-only sensitivity is severely underpowered (n = 3 ALS); a null result there is uninformative, but a positive result is informative. The age-adjusted regression assumes a linear age effect and may underestimate confounding for features with non-linear age dependence.
